# Socio-economic and environmental patterns behind H1N1 spreading in Sweden

**DOI:** 10.1101/2020.03.18.20038349

**Authors:** András Bóta, Martin Holmberg, Lauren Gardner, Martin Rosvall

## Abstract

The first influenza pandemic in our century started in 2009, spreading from Mexico to the rest of the world, infecting a noticeable fraction of the world population. The outbreak reached Europe in late April, and eventually, almost all countries had confirmed H1N1 cases. On 6 May, Swedish authorities reported the first confirmed influenza case. By the time the pandemic ended, more than 10 thousand people were infected in the country. In this paper, we aim to discover critical socio-economic, travel, and environmental factors contributing to the spreading of H1N1 in Sweden covering six years between 2009 and 2015, focusing on 1. the onset and 2. the peak of the epidemic phase in each municipality.

We apply the Generalized Inverse Infection Method (GIIM) to identify these factors. GIIM represents an epidemic spreading process on a network of nodes corresponding to geographical objects, connected by links indicating travel routes, and transmission probabilities assigned to the links guiding the infection process. The GIIM method uses observations on a real-life outbreak as a training dataset to estimate these probabilities and construct a simulated outbreak matching the training data as close as possible.

Our results show that the influenza outbreaks considered in this study are mainly driven by the largest population centers in the country. Also, changes in temperature have a noticeable effect. Other socio-economic factors contribute only moderately to the epidemic peak and have a negligible effect on the epidemic onset. We also demonstrate that by training our model on the 2009 outbreak, we can predict the timing of the epidemic onset in the following five seasons with good accuracy.

The model proposed in this paper provides a real-time decision support tool advising on resource allocation and surveillance. Furthermore, while this study only considers H1N1 outbreaks, the model can be adapted to other influenza strains or diseases with a similar transmission mechanism.

## I. Introduction

A novel pandemic emerged in Mexico during the spring of 2009, caused by a recombined influenza strain derived from circulating swine influenza strains. It spread quickly to other continents, and struck Europe with a spring/summer wave, affecting several countries. The rate of transmission subsided as the summer progressed but accelerated again during the autumn season, this time in all European countries. The spread followed a west to east progression, a typical pattern for seasonal influenza [24].

The first cases in Sweden appeared late in April, and local transmission followed, mainly in the major cities. In late September-early October, a nationwide outbreak started, peaking in November. From mid-May, the Swedish institute for infectious disease control required mandatory reporting of individual cases, which later changed to hospitalized patients supplemented by laboratory notification of positive samples [25]. WHO declared the end of the pandemic on 10 August 2010. However, the mandatory reporting of the H1N1 pandemic strain continued in Sweden during the following five seasons, providing a detailed and thorough collection of epidemic data on a fine spatial resolution.

The spatial component of many infectious diseases is crucial for the understanding of epidemic transmission. Recent years have seen a surge in the use of mathematical models to describe the geographic transmission of infectious diseases [6], [19]. Specifically, with the increasing availability of geotagged epidemiological data, the 2009 A(H1N1) pandemic influenza has been studied using spatially explicit models. For example, studies in the US have revealed human mobility patterns as important mechanisms for influenza epidemic transmission [7], [10], [14]. A previous geographic study of the pandemic spread over Sweden indicated a progression from the north to the south during the year 2009 [21]. However, the significant risk factors that drive the influenza dynamics in Sweden have not been studied in detail.

Meteorological factors associated with the rate of influenza transmission among individuals include precipitation, humidity, temperature, and sun radiation[5], [18], [8]. However, uncertainties in the data remain, and meteorological drivers may play more significant roles in some geographic regions than others. The role of population size and density in cities is also known to affect the dynamics of influenza transmission and the weight of environmental factors [8]. Apart from the importance of schoolchildren for influenza transmission [27], socio-economic factors important for influenza spread remain sparsely studied [16]. Moreover, the relative contributions of multiple risk factors that drive the spread of influenza are poorly understood, underlining the need for new methods and models to manage the increasing volume of spatio-temporal disease data and socio-economic and environmental metadata coupled to these data.

In this paper, we seek to uncover the critical socioeconomic, travel, and environmental factors behind H1N1 outbreaks in Sweden, covering six years between 2009 and 2015. We will focus on two properties of the outbreaks: the start and the peak of the epidemic phase. We aim to identify the relationship between the timing of these events and the above factors available for each of the 290 municipalities of Sweden. We also explore the predictive capabilities of our approach by using the 2009 pandemic to train our model and measuring its performance on data from the following years. We use a network model to define the paths of disease spreading between the municipalities. In this model, each municipality defines a node, and municipalities are connected if there is significant travel between them based on a travel survey covering multiple years. We use the Generalized Inverse Infection Model (GIIM) [2], [3], [9] to establish the relationship between the properties of the outbreak and several socio-economic, travel, and environmental indicators. GIIM is a network-based optimization method that seeks to estimate the transmission risk between geographical areas by a risk function of known attributes. The risk function assigns a weight to each attribute representing their relative contribution. The method estimates the parameters of this function through iterative refinement by repeatedly running a simulated infection process. An error function measuring the difference between the output of the simulated process and an observed real-life outbreak guides the optimization process, which GIIM solves with a Fully Informed Particle Swarm Optimization Method [11].

According to our results, the most critical factor that guides the spread of H1N1 in the municipalities of Sweden is population size, followed by temperature. GIIM also estimates the likelihood of disease spreading between the municipalities, and the importation and exportation risks for all municipalities. More importantly, we can use the risk factors, their assigned weights, and the attribute function to simulate an outbreak at any given period, allowing us to predict the timing of epidemic events in future seasons. By training our model on data from the 2009 pandemic, we can predict the timing of the above epidemic events in later seasons with good accuracy.

## II. Data

Our analysis covers Sweden on the geographical level of its 290 municipalities. Almost all the data sources are domestic, and are available either on the municipality level (only the cities of Stockholm and Gothenburg comprise more than one municipality), or the DeSo (demographic statistical areas) [17] level, a subdivision of the municipality level. The only exception is the weather-station based climate data, which we have converted from specific geographic coordinates to the municipality level. Because Sweden covers a large geographical area with significant variance in features, climate, and population, the selected indicators may vary significantly.

### A. Maps and geographical data

We constructed the maps of Sweden and its municipalities from open-source shapefiles obtained from [17] using the QGIS software [23].

### B. Travel Data

Travel patterns in the Swedish population have been recorded since the mid-1990s by interviewing representative selections of several thousand 6-84-year-old inhabitants. As part of this study, we obtained complete questionnaire data for the years 2011-16 [26]. The data contains posts on the commuting habits of individuals on a daily level with partial and whole trips and the geographic origin and target of the travels at the DeSo level. Since DeSos are subdivisions of municipalities, we converted them to the municipality level. The survey contains additional information, including the age and income of participants and the means of travel. However, we omitted this information from the analysis because we gathered more detailed information from the Swedish statistical bureau.

Since this study considers the years from 2009 to 2015, we have used the 2011-2016 period to define the travel patterns for all parts of the analysis, including the years 2009 and 2010. We have no reason to assume that travel patterns changed significantly between the years 2009 and 2016. Furthermore, while the sample size of the survey is adequate to calculate general statistics, it is too sparse to construct a travel network with temporal dynamics, even on a yearly level. Instead, we constructed an aggregated static network containing all feasible routes of infection. To partially compensate for the inadequacies of the survey, we made sure that the travel network represents all air and rail travel routes and connects all neighboring municipalities. The sparsity of the travel survey also prevents us from reliably measuring the frequency of travel between the municipalities. To include at least one travel-related variable, we selected node degree – the number of travel routes connecting a municipality to other municipalities – to signal the importance of a municipality in the travel network of Sweden.

### C. Epidemiological data

Our data set consists of all laboratory-verified cases of A(H1N1)pdm between May 2009 and December 2015, extracted from the SmiNet register of notifiable diseases, held by the Public Health Agency of Sweden. While the number of flu cases is regularly underreported, the SmiNet database contains 16000 records, which is a reasonably large sample compared to other notifiable diseases. Due to confidentiality reasons, cases are anonymized, and addresses are aggregated at the DeSo level together with the date of diagnosis, age, and gender. To make sure the addresses represent the habitation at the time of diagnosis, the register was cross-referenced at Statistics Sweden (SCB) with a historical address register before anonymization. We obtained ethical approval for the data acquisition.

Consequently, the epidemiological data contains the number of daily reported cases at the DeSo level, which we converted to the municipality level. Since the available data covers an extended period from 2009 to 2015, we partitioned the case counts into six separate outbreaks corresponding to the 2009/2010, 2010/2011, 2011/2012, 2012/2013, 2013/2014, and 2014/2015 flu seasons. The size and timing of the outbreaks show considerable differences. The 09/10 swine flu pandemic covered a significant part of the year 2009 and lasted into the first weeks of 2010. In contrast, the 11/12 season saw almost no flu cases. Figure 2/B shows the number of new cases in 2009 from week 37 to 50 in a few selected municipalities.

### D. Socio-economic data

One of the main goals of this paper is to establish a relationship between socio-economic factors and epidemic spreading. Since this study only considers the country of Sweden, high-quality data is readily available from Statistics Sweden [22], the organization responsible for coordinating the system for the official statistics in the country. Based on available statistics and previous studies [9], [27], [16], [8], we selected: 1. The average household income as an economic indicator. 2. The average number of children younger than 21 years per household to indicate family size. 3. The number of people receiving social aid to represent poverty in a region. 4. Population size and population density as the number of people per sq. km of land area. All the above statistics are available at the municipality level. Figure 1 shows the geographical distribution of population size and the average number of children per household.

**Fig. 1.**
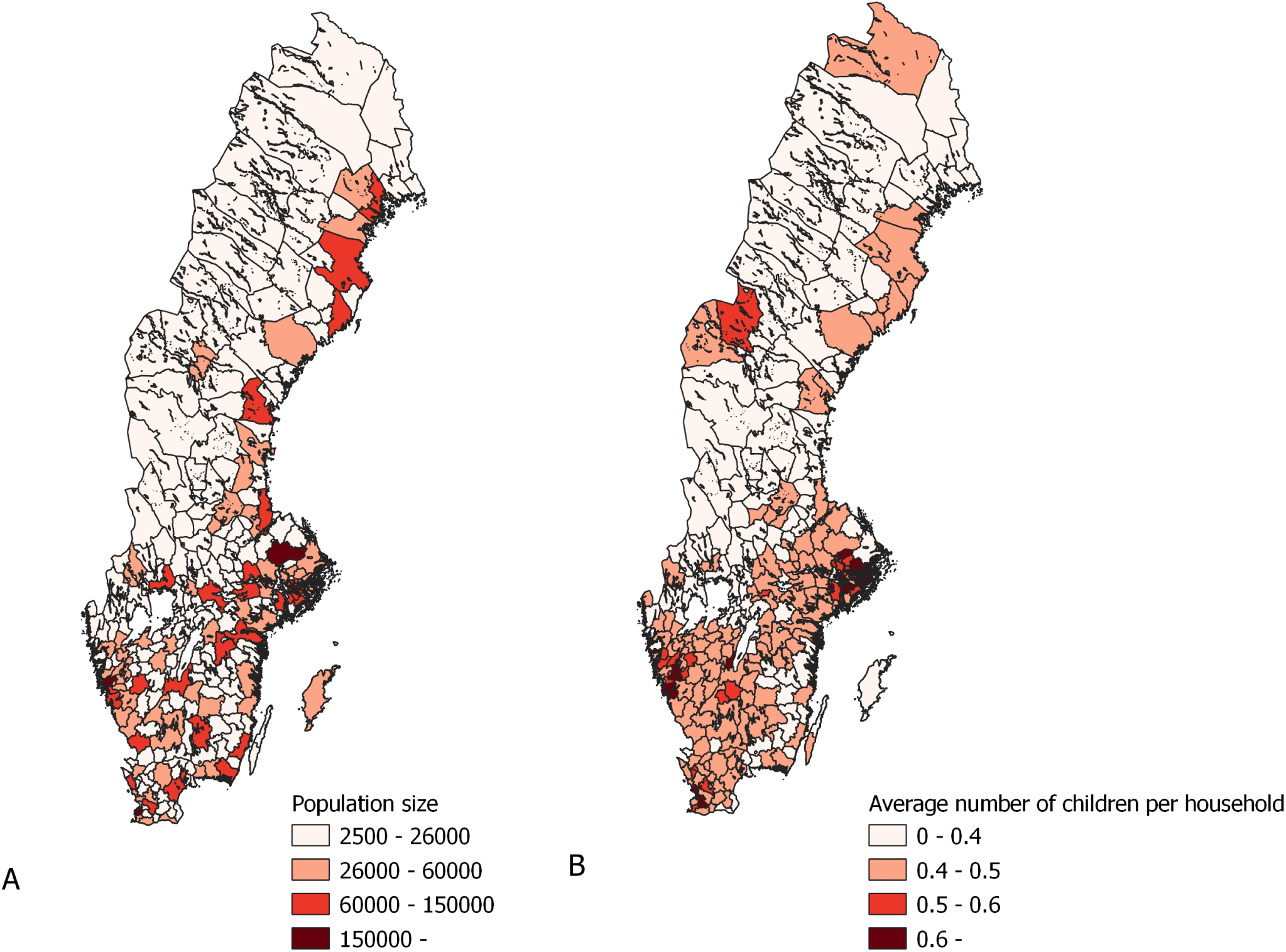
Geographical distribution of population size and the average number of children per household of the municipalities of Sweden.

Both population density and population size have been used before to model epidemic spreading [8]. In a geographical setting population density is preferred due to its independence from inequalities in the size of geographical areas. In contrast, using population size can be misleading if differences are more than minimal. However, the fine geographical resolution of the municipalities of Sweden and the lack of huge population centers (except for Stockholm and Gothenburg) allows us to use population size as a variable. We experimented with both density and size in our study. We found that density only has a small contribution to the timing of the epidemic events, while population size is a significant factor. Therefore, we chose to include population size.

### E. Climate data

Even though the exact mechanisms are unknown, the relationship between environmental temperature and humidity and seasonal influenza is well established in the literature [20], [5], [18], [8]. As such, it is one of the key factors used in this study. Due to Sweden’s geographical position and the effect of the Gulf Stream, the country’s climate ranges from an oceanic climate in the far south to a subarctic climate in the far north, while central Sweden has humid continental climate.

We obtained detailed climate data from the European Climate Assessment Dataset [15]. This database contains daily meteorological station observations covering Europe. Of the elements available in the database, we included mean temperature and relative humidity as factors in this study. Using the geographical coordinates of the weather stations and the shapefiles of Sweden and its municipalities, we assigned each weather station to its municipality, then averaged the readings inside municipalities in case there were multiple stations assigned to them. If a municipality did not have an assigned meteorological station, we used an average of the values from neighboring municipalities. We show the averaged mean temperatures of a few select municipalities in Figure 2/C.

**Fig. 2.**
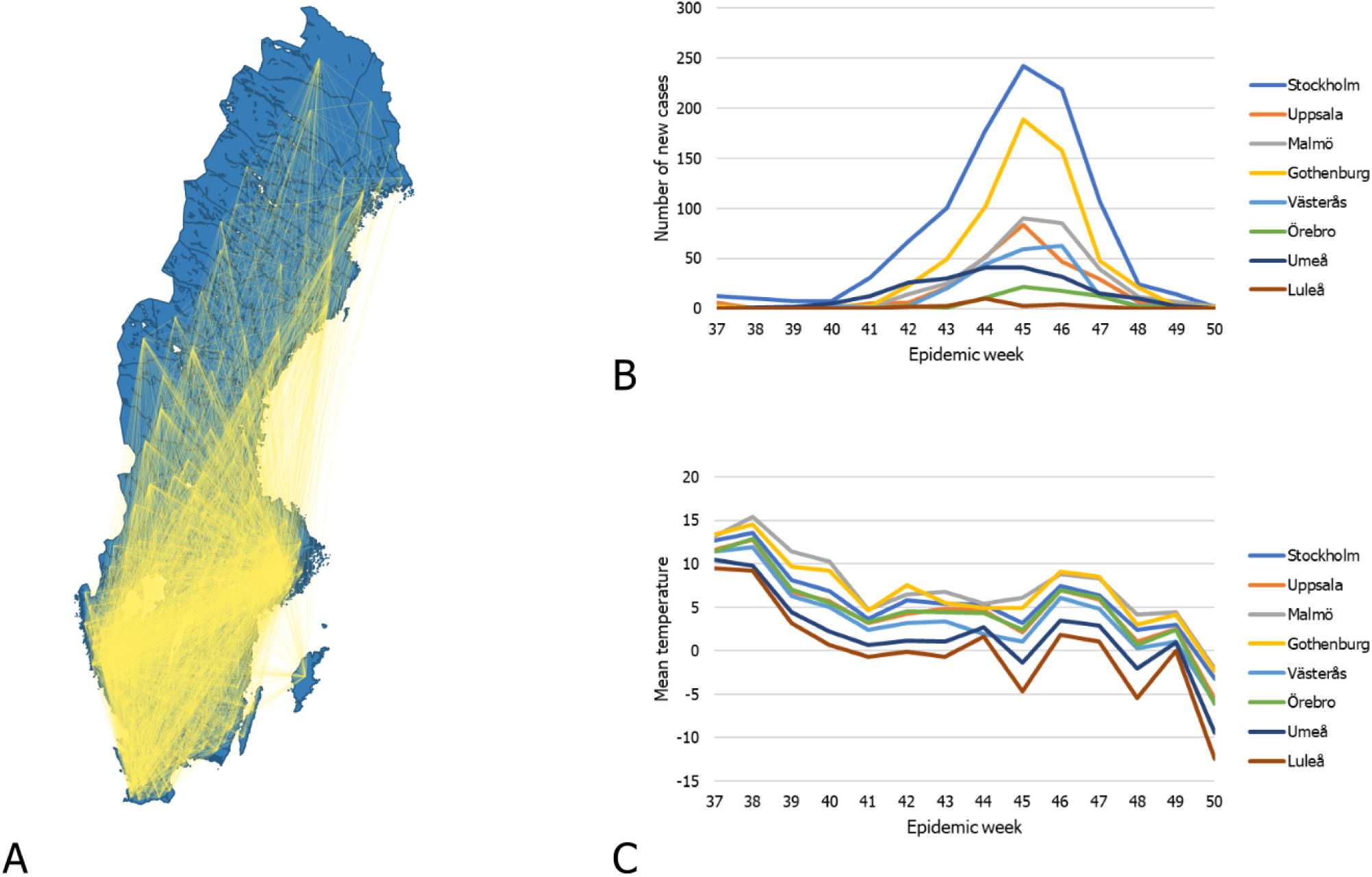
A. The travel network of Sweden. B. The number of new cases in from epidemic week 37 to 50 in the municipalities listed on the right side. C. The average mean temperature from week 37 to 50 in the municipalities listed on the right side.

## III. Method

We use the Generalized Inverse Infection Model (GIIM) [3] to identify the relationship between the H1N1 outbreaks and the socio-economic and climate factors. The GIIM method is a network optimization tool for infection processes, which has been successfully applied to real-world problems [4], including the modeling of the geographical spread and transmission of Zika in the Americas [9]. GIIM brings a novel approach to epidemic modeling because instead of merely simulating an outbreak, the method estimates the parameters and properties of an infection process using observations from an actual outbreak.

The GIIM method uses a network to represent the geographical areas involved in the outbreak as nodes, and allows the assignment of multiple attributes to the nodes and links of the network. The method also relies on observations from an actual outbreak with weekly case counts reported in geographical areas. In practice, GIIM uses single properties of the epidemic curve, such as the maximum case count or the onset of the outbreak. GIIM uses an iterative refinement approach to reconstruct the outbreak from the network and its assigned attributes by matching the properties of a simulated outbreak to the observed properties of the actual outbreak, *i*.*e*. by minimizing the error between the estimated and observed data. The method identifies the transmission risks on the links of the network defined as a function of the known attributes, and returns the parameters of this transmission risk function.

### A. Inputs

The GIIM method requires three inputs: an underlying network structure, attributes assigned to the nodes and edges of the network, and a set of observations on a real-life transmission process. The underlying network structure in this study represents the municipalities of Sweden. We denote graph *G* as *G*(*V, E*), where *V*_*G*_ is a set containing all the vertices of the graph, while *E*_*G*_ contains all the edges of the graph. We denote the edges of *G* as *e*_*u,v*_ ∈ *E*_*G*_ with *u* as the tail and *v* as the head of the edge, where *u* and *v* are nodes of the network and edge *e*_*u,v*_ links node *u* to *v*. The nodes of the network represent the 290 municipalities of Sweden, and we define the edges of the network based on the travel survey introduced in the data section. The survey contains travel information for all participants indicating the source and target municipalities of their travels between 2011 and 2016. We created a directed edge between two municipalities A and B if at least one individual traveled from A to B. To ensure that we represented all feasible travel paths in the network, we connected all neighboring municipalities. In this way, the edges of the network indicate significant recorded travel between the pair of municipalities they connect. We denote this network *G*_*S*_ and illustrate it in Figure 2/A.

#### Attributes

We represent the previously defined socioeconomic, travel, and climate factors as attributes assigned to the nodes of the network. All attributes are real values normalized between zero and one. We used the following attributes:

1. 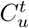 the incidence of new flu cases reported at municipality *u* in week *t*.
2. *D*_*u*_ the degree of node *u*. Lacking more accurate travel indicators, we used this value to represent the importance of a municipality in the travel network of Sweden.
3. 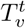 the mean temperature measured in municipality *v* in week *t*.
4. 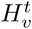 the absolute humidity measured in municipality *v* in week *t*.
5. *P*_*v*_ the population size of municipality *v*.
6. *I*_*v*_ the average income per household in municipality *v*.
7. *S*_*v*_ the number of people receiving social aid in municipality *v*.
8. *K*_*v*_ the number of children under 18 years of age per household in municipality *v*.

Attributes marked with a time index *t* are dynamic. As such, their values change in time depending on the week of the outbreak. We build our model based on the 2009 flu pandemic. Therefore, all dynamic attributes refer to the 2009/2010 season unless noted otherwise. We only use data from the 2010-2015 period in the last part of our analysis, when we test the predictive ability of our model.

#### Attribute function

To reconstruct the observed outbreak using the network and its attributes, GIIM repeatedly runs a simulated infection process. The infection process requires transmission probabilities (also called edge infection probabilities) 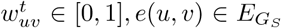 assigned to the edges of the network. The flexibility of the GIIM model allows us to define these values as a function of known attributes, and in this paper we define the functions with the attributes listed above.

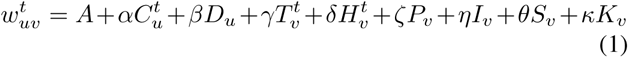

The variables in equation (1) cover the attributes listed previously, and the optimization algorithm of GIIM estimates the coefficients of these functions.

#### Reference outbreak

The GIIM method seeks to estimate the parameters of an existing outbreak. Reference observations provide information about this outbreak: a set of vectors, each containing an indicator value on the state of the infection process at all nodes. Multiple vectors provide information at different time periods of the process. While observations can take different forms [3], here we define them as a set of binary vectors covering a selected set of weeks from each of the yearly outbreaks covered in this study. Each binary vector corresponds to a week, and a value of 1 indicates that the epidemic peak or onset already happened, while a 0 indicates that it has not happened yet. The week of the peak or onset has a value of 1. We aim to estimate two different parameters of the outbreak: 1. the timing of the epidemic peak, *i*.*e*. the week with the highest number of newly infected cases and 2. the timing of the epidemic onset, the week when the number of newly infected cases increases significantly compared to the previous period.

The timing of the epidemic seasons varies depending on the year. Table 1 lists the weeks in each season when there was a significant number of flu cases. We used these weeks to construct the reference observations. The 2009-2010 season had flu cases all over the year, but we focus on the fall of 2009, where the largest portion of the outbreak took place.

**TABLE I.** The start and end week of each epidemic season.

| Season | Start week | End week |
| --- | --- | --- |
| 2009-2010 | 2009 week 37 | 2009 week 50 |
| 2010-2011 | 2010 week 50 | 2011 week 10 |
| 2011-2012 | 2011 week 51 | 2012 week 09 |
| 2012-2013 | 2012 week 51 | 2013 week 10 |
| 2013-2014 | 2013 week 49 | 2014 week 11 |
| 2014-2015 | 2015 week 5 | 2015 week 14 |

### B. Infection Model

As part of its optimization process, GIIM relies on the repeated evaluation of a simulated infection process. In this study, we adopt the SI compartmental infection model defined for networks [3], [12], which was successfully used in a similar study [9]. Part of the more general SEIR infection model family, the SI model only has two states: susceptible (S) and infected (I), representing infectious nodes, which continuously try to infect their healthy neighbors, and susceptible nodes prone to infection. Each node of the network has a state during the process, which may change over time. We assign edge infection probabilities 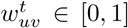 to all edges of the network. The *t* time index indicates that these probabilities may change their values depending on the discrete time scale of the process. However, 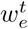 is strictly an input of the model and does not depend on the spreading process in any way. The infection process is iterative and takes place in a finite number of discrete time steps. In each iteration, a node may change its state depending on the state of its neighbors and the edge infection probabilities assigned to the edges connecting it to them. Nodes may change their states from susceptible to infected, but infected nodes stay infected until the end of the process. The total number of discrete time steps the process takes is limited to the number of weeks with reported new flu cases.

As in [3], [9], we define the SI infection model using a graph *G*(*V, E*) with edge infection probabilities 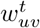 assigned to all of its edges and the initial set of infected nodes *A*_0_. The rest of the nodes are in the susceptible state at the beginning of the process. Let *A*_*t*_ *⊆ V*_*G*_ be the set of infected nodes in iteration *t*. In each iteration *t* each infected node *u* ∈ *A*_*t*_ tries to infect all its susceptible neighbors *v* ∈ *V*_*G*_ | *A*_*t*_ depending on the edge infection probability 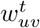 of the edge connecting them. If the attempt is successful, *υ* joins the set of infected nodes in the following iteration. If more than one node is trying to infect *υ* in the same iteration, the attempts are made independently of each other in an arbitrary order within the same iteration. The process terminates naturally if all nodes reachable from the initially infected nodes with nonzero edge infection probabilities adopt the infected state, or when there are no more reported new flu cases.

The above process defines a single instance of an outbreak. Instead of binary values, GIIM requires for each node the likelihood of being in an infectious state for all time steps. Therefore, we ran the process 5000 times and counted the number of instances when the nodes of the network were infected [13], [1]. When we refer to the output of the infection model, we refer to the estimated likelihood values as opposed to the binary outputs of a single instance.

### C. The GIIM Method

The GIIM method [3] defines the problem of estimating edge infection probabilities as an optimization task. Its inputs include a network, several attributes on the nodes and the edges of the network, and a set of reference observations of an actual outbreak. GIIM provides an estimation of the observed outbreak by simulating one. Apart from the result of the simulation (which may be more detailed than the original), its output provides an assignment of edge infection probabilities and the relative importance of the attributes according to equation (1).

To define GIIM’s inputs, let 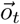 denote a vector containing observations on an infection process. Let 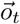 assign a value to all *υ* ∈ *V*_*G*_. Let *t* ∈ *T* denote a discrete time stamp indicating the week in which the observation was taken, and *T* be the set of all time stamps. Let *O* denote the set of all observations 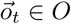 for all *t* ∈ *T*. Let *ℐ* denote the SI infection model introduced in the previous subsection, and *W*_*G*_ : *E*_*G*_ *→* [0, 1] be the initially unknown assignment of edge weights to the edges of the graph. Finally, let *Inf* be a procedure, which makes observations on infection process *ℐ* at sample times *T*, taking place on graph *G* with assigned edge weights *w*_*e*_ ∈ *W, e* ∈ *E*(*G*). We denote *Inf* as *O* = *Inf* (*G, W, ℐ, T*).

#### General Inverse Infection Model

*Given an unweighted graph G, infection model ℐ, the set of sample times T, and reference observations O* = *Inf* (*G, W, ℐ, T*), *we seek the edge infection probability assignment W*^′^ *such that the difference between O and O*′ = *Inf* (*G, W*^′^, *ℐ, T*) *is minimal*.

In this study the set of reference observations *O* contains binary vectors indicating the timing of the epidemic onset and peak, while the observations *O*^*’*^ generated by running the infection model are real-valued. To compute the difference between *O* and *O*^*’*^, we employ ROC evaluation. We pairwise compare vectors 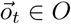 and 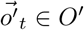 for all *t* ∈ *T*, calculating the AUC value for each pair and averaging over all pairs. The GIIM method uses an iterative refinement algorithm to solve the optimization task above. Starting from an initially random edge weight assignment, GIIM simulates an outbreak and compares the output with reference observations. Then it updates the edge weight assignments and repeats the process. The search algorithm uses the Fully Informed Particle Swarm Optimization method [11], a multi-agent iterative optimization algorithm, which was previously shown to perform well with GIIM [3], [9]. Algorithm 1 [9] summarizes the iterative GIIM algorithm.

##### Algorithm 1 Generalized Inverse Infection Model

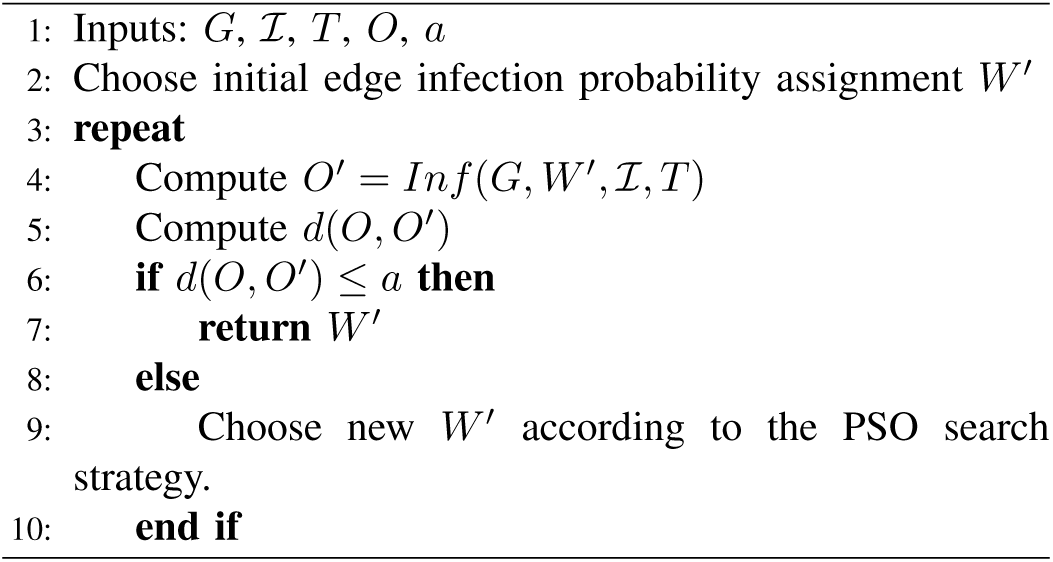

Estimating individual edges is difficult due to the size of most networks. To avoid this problem, GIIM defines the edge weights as a function of known attributes on the nodes or edges of the graph. This way, the goal of the optimization task is to find the coefficients of this function. It is also possible to define dynamic attributes or edge weights, indicating that their value changes in time. In practice, this means that the value of the edge weight or attribute is a function of *t*, a discrete time stamp corresponding to the actual iteration of the infection model.

The general form of the edge function can be written as 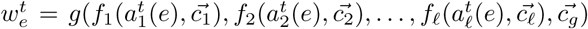 for all *e* 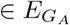, where 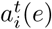 represents the *i*-th attribute on edge *e* ∈ *E*_*G*_ at iteration *t* of the infection process, *l* denotes the number of available attributes, *f*_1_, …, *f*_*l*_ and *g* are functions and 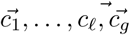, are coefficients of functions *f*_*1*_, *…, f*_*l*_, *g*. This formulation is easy to implement and allows us to assign different functions to different attributes, while the role of function *g* is to aggregate and normalize the results of the individual attribute functions to ensure they fall between 0 and 1. The value 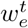 denotes the edge weights and *C* the set of all coefficient vectors.

Using the function-based alternative greatly simplifies the optimization task, reducing the number of values we have to estimate from |*W*| = |*E*_*G*_| to |*C*|. However, its main advantage is that instead of providing a single individual value for each edge, it allows us to explore the relationship between the factors potentially related to the outbreak and the outbreak itself. Equation (1) defines the edge functions used in this study. We trim the weighted sums above 1 and below 0 by taking 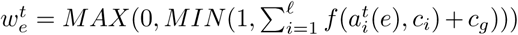. To reduce the solution space of the PSO method and to make the results of individual test runs comparable, we bound all parameters between −0.5 and 0.5, except *A*, which we bound between −1 and 1.

## IV Results and Discussion

We start our analysis by studying the spreading of H1N1 in the autumn of 2009, weeks 37–50 for the epidemic onset and 39–50 for the peak, corresponding to a period from mid-September to mid-December. We model the outbreak on a weekly basis and set up the input files of GIIM as described in the Inputs section, To partially account for the summer wave of infections in 2009, we select the municipalities of Stockholm, Malmo and Gothenburg as infected in the starting week of the analysis, as these municipalities contained most infected individuals during the summer wave. We use the attribute function defined in equation (1) with the socio-economic, travel and environmental indicators introduced in the Inputs section. To compensate for the stochastic nature of the model, we ran the algorithm 20 times with the same set of inputs and computed the mean and variance of the results.

### A. Model accuracy

We evaluated the accuracy of our model by independently computing the ROC AUC value for each week of the estimation process, 14 weeks for the onset estimation and 12 weeks for the peak estimation. The averaged AUC value for onset estimation was 0.875, indicating a good fit of our model, while the averaged AUC value of peak estimation was 0.817, which indicates a moderately accurate fit. Figure 3 shows individual AUC values for all weeks for both objectives of the estimation. The estimation is easy during the first weeks due to the small number of positive examples. In the majority of the municipalities, the fall H1N1 outbreak started and peaked in weeks 42–45, where the accuracy of the estimation process drops. Onset estimation accuracy around 0.82 for these critical weeks indicates some uncertainty in the exact timing of the onset, but most estimates indicate good accuracy. Peak estimation is significantly less accurate, around and below 0.7 for two weeks, and stabilizing around 0.78 for the rest of the weeks. In summary, while epidemic peaks are more difficult to estimate, onset estimation has good accuracy for the entire observation period except for slight uncertainty around the most eventful weeks (Figure 3).

**Fig. 3.**
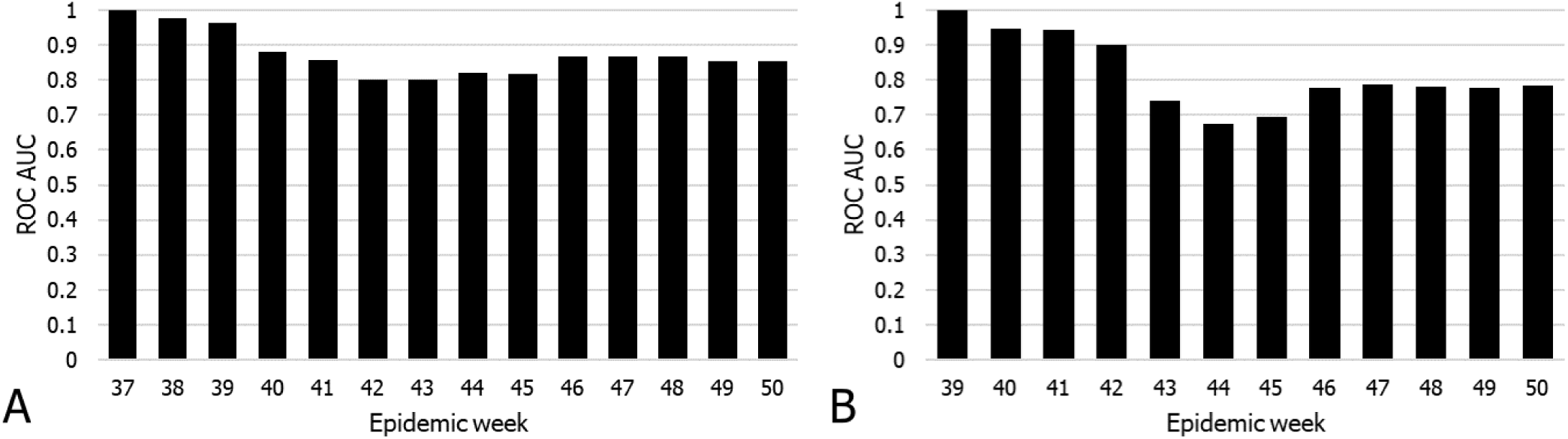
Model accuracy (ROC AUC) for all weeks of the observation period. A. Onset estimation. B. Peak Estimation.

### B. Risk factors

To evaluate the relative contributions of the travel, socio-economic, and climate factors on the timing of the epidemic onset and peak in the municipalities of Sweden, we experimented with multiple combinations and functions of them. We found that equation (1) provides the best fit to our reference outbreak. Figure 4 shows the mean and standard deviation of the coefficients assigned to the risk factors, while equations 2 and 3 show the attribute function with the mean of the estimated coefficients for epidemic onset and peak estimation respectively.

**Fig. 4.**
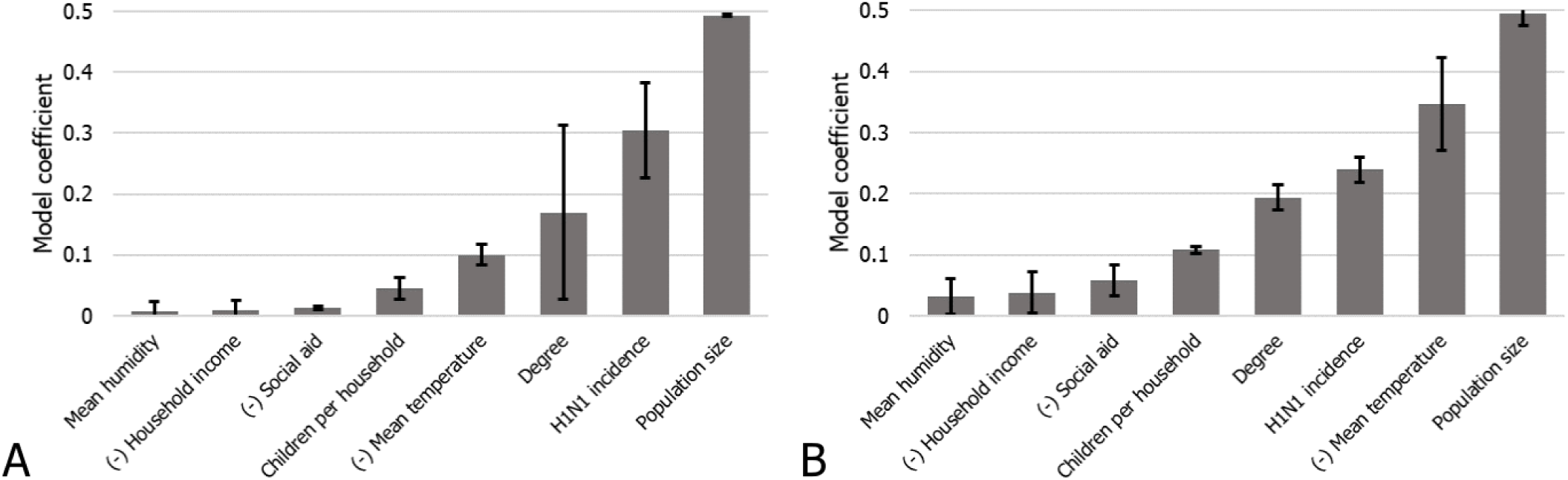
Mean and standard deviation of the coefficients assigned to the risk factors by GIIM. A. Onset estimation. B. Peak Estimation.

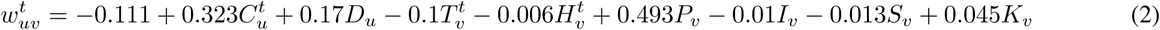

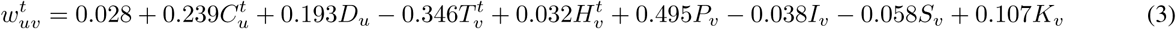

The weights in equations 2 and 3 represent the relative contribution of each risk factor to the transmission risk between the municipalities. The standard deviation of the coefficients across test runs is small except for node degree in onset estimation. More importantly, the relative importance of the risk factors remains robust.

The coefficients for onset and peak estimation show some similarities, but there are differences. The most significant factor in the timing of the epidemic onset and peak is the population size of the municipalities, indicating that the disease reaches and peaks in large population centers first and then spreads to the countryside. Accordingly, node degree, representing the importance of a municipality in the travel network of Sweden, receives a moderately positive coefficient. A significant factor in onset estimation is the incidence of H1N1 cases in the municipality on the source of the links. The same coefficient gets a much smaller weight in peak estimation since the virus is already present in the target municipality. This phenomenon highlights the difference between estimation goals.

Although the relationship is not direct, environmental factors such as temperature, play a critical role in the appearance of influenza. According to our model, the mean temperature contributes slightly to the onset of the disease with a small negative coefficient. At the same time, the temperature is the second most significant risk factor to peak estimation, where it has a large negative weight. This result implies that instead of heralding the beginning of the flu season, a drop in temperature greatly aggravates the effect of a current outbreak, driving it to peak intensity. We also included humidity in our model, but in both estimations, it only provides a small contribution to the spreading risk.

Perhaps due to the relative homogeneity of a single country, we found that the effect of the socio-economic variables on the transmission risk between municipalities to be minimal. The average number of school-age children per household, which is a well-known risk factor [27], showed the most significant influence. Like mean temperature, it contributes more to the timing of epidemic peaks than to the start of an outbreak, and even for peaks, it only has a low-moderate coefficient. The influence of income is close to zero in both estimation types, whereas the number of people receiving social aid adds a small negative contribution to peak estimation.

We conclude that different factors contribute to the onset and peak of H1N1 outbreaks in the municipalities of Sweden during the 2009 flu pandemic. The timing of the epidemic onset is mainly influenced by population size, the position of the municipality in the travel network and the number of cases in neighboring municipalities. This result is in line with the spreading mechanism of more traditional agent-based models where the outbreak travels from region to region after infecting a certain amount of people locally, favoring population centers. Temperature has a small negative effect, while school-aged children have a small positive effect. In contrast, apart from population size, the timing of the peak highly depends on mean temperature, and other socio-economic factors such as schoolaged children and social aid have a more pronounced effect too. This result implies that the timing of the peak depends more on local risk factors, as opposed to the more network-oriented spreading of the epidemic onset.

### C. Exportation, importation and route level risk

The edge infection or transmission probabilities between the municipalities of Sweden are among the outputs of GIIM. In addition, it is possible to define a node-based import and export risk weight by aggregating the edge-based risk values on the in- and out-edges of each node. This value is also known as node strength. It is important to note, that these values are not probabilities, but risk indicators of importing or exporting the disease from or to a municipality. They represent a relative weight that allows us to rank the municipalities.

Figure 5/A shows the geographical distribution of exportation risk at week 41, and Figure 6/A illustrates the time-dependent exportation risk values for some of the highest risk municipalities in onset estimation (for a complete ranking, see the supplementary material). Large population centers appear at the top of the list. While Gothenburg and Stockholm remain at the top, Malmö only stays in the top 20 exporters during the initial weeks, with its relevance decreasing even more later in the outbreak. Other larger cities include Linköping, Jönköping, and Uppsala, although except for Uppsala, their relevance fades in time. Several of the larger cities of northern Sweden, such as Skellefteå, Umeå, and Östersund, are top exporters until mid-November, partially confirming the observations in [21].

**Fig. 5.**
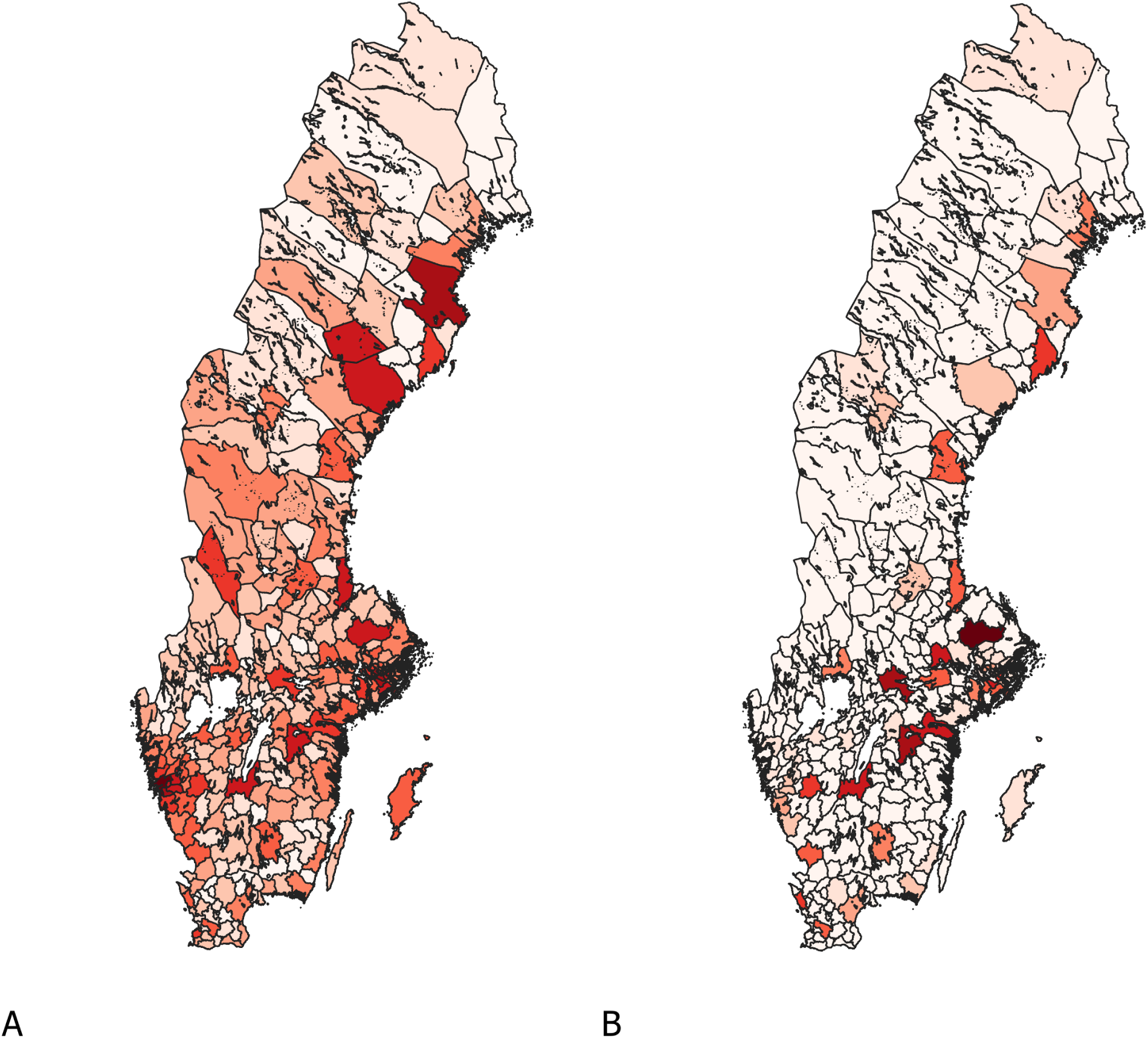
Geographical distribution of exportation (A) and importation (B) risk at week 41 in onset estimation.

**Fig. 6.**
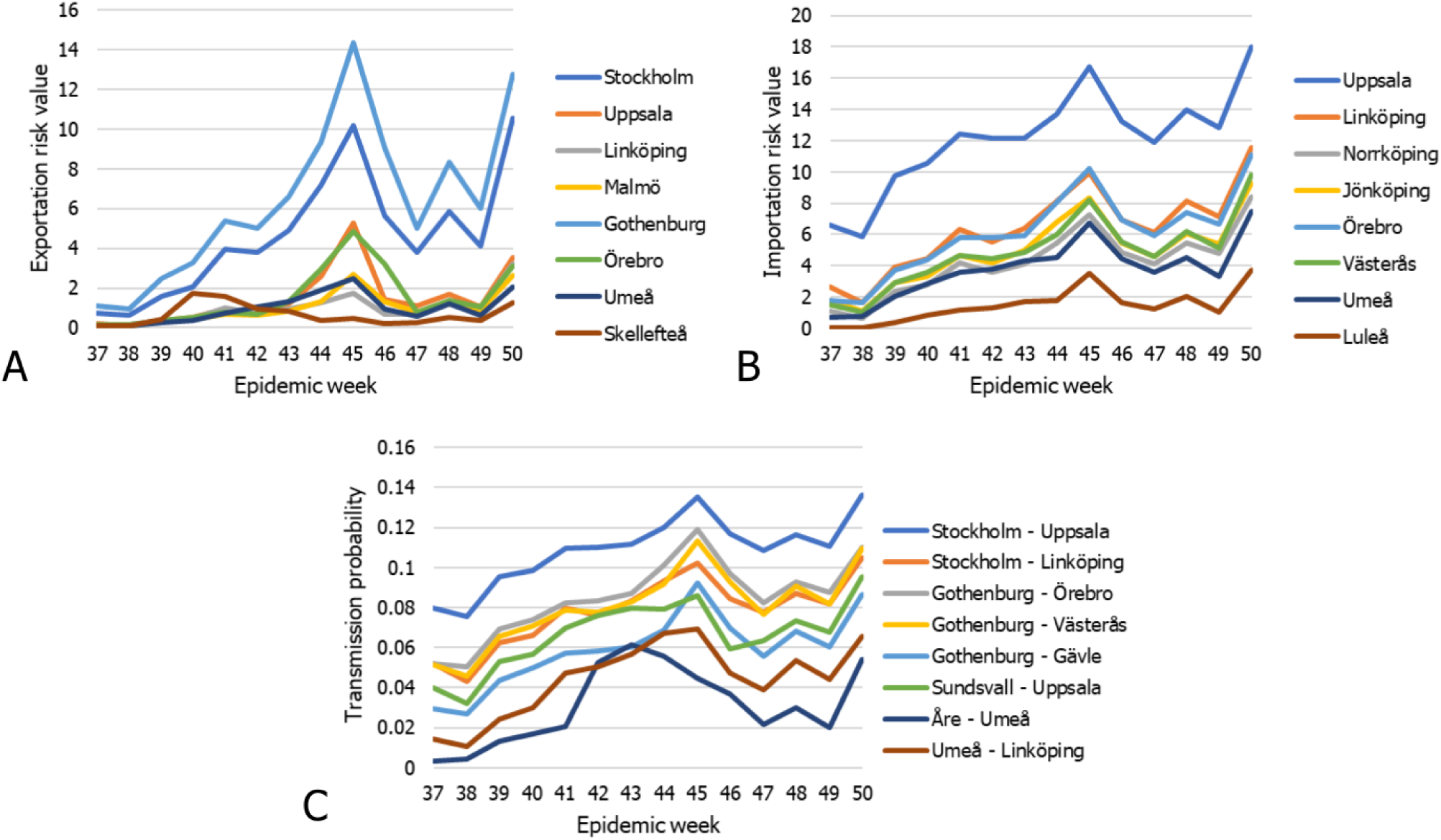
A. Exportation risk values of select municipalities in onset estimation. B. Importation risk values of select municipalities in onset estimation. C. Transmission probabilities of select links in onset estimation.

Figure 5/B shows the geographical distribution of importation risk at week 41, while Figure 6/B illustrates the time-dependent importation risk values for some high-risk municipalities in onset estimation (for a complete ranking, see the supplementary material). This ranking is more static than the export ranking, with large population centers remaining high risk until the end of the outbreak. These cities include Linköping, Jönköping, Norrköping, Uppsala, Örebro and Västerås, together with a few northern municipalities such as Umeå, Luleå and Sundsvall. In contrast to the export risk ranking, Stockholm county and Västra Götaland are not top-import municipalities.

Due to a large number of edges in the graph, it is difficult to highlight the individual links most important to the outbreak. Figure 6/E shows some of these links. For example, early in the outbreak, links connecting to Uppsala appear most prominently among links to Umeå, Linköping, Örebro and Västerås. This pattern remains until the end of the outbreak. and Central Sweden and moving north along the coast of the outbreak.

Starting from mid-October, links connecting the municipalities of Northern Sweden appear at the top of the edge-based risk ranking and stay there for several weeks until mid-November. At the end of our observation period, links connecting larger population centers in South and Central Sweden dominate the top of the ranking.

Our observations on the timing of the epidemic onset of H1N1 in the fall of 2009 follow a loose geographical pattern. Following the smaller summer wave of infections, the disease stays in the largest cities of the country, and from September begins slowly spreading to the other larger population centers of South and Central Sweden and a few cities in Northern Sweden. A massive burst of the outbreak happens in mid-October, reaching the countryside in South and Central Sweden and moving north along the coast of the Gulf of Bothnia infecting the larger population centers on its way. After peaking in most of the country in early and mid-November, the outbreak first dies in the north and finally in the largest cities where it began.

### D. Model predictability

To estimate parameters of future H1N1 outbreaks in Sweden, we focus on the timing on the epidemic peak and the epidemic onset in the flu outbreak in epidemic seasons 10/11, 12/13, 13/14, 14/15. We omitted season 11/12 from our analysis due to the small number of H1N1 cases in the country in this season, possibly an effect of the extensive mass vaccination with a pandemic vaccine during 2009. We set up the reference observations for our four target seasons in the same way as for the 09/10 season (for observation periods of these seasons, see Table 1). We also updated our dynamic input attributes to correspond to the actual temperature and humidity values in the same time periods. Finally, we defined the infection sources for each of the target seasons and estimation types by selecting the three municipalities with the earliest onset and peak, respectively.

We used the estimated function coefficients for the 09/10 season as defined in equations (2) and (3) and the corresponding inputs to run an instance of the infection model for each of our four target seasons. We compared this output with the reference observations for each of the target seasons by computing the ROC AUC metric. There are considerable differences between the predictive abilities of the two estimation types (Figures 7 and 8). Onset estimation performs significantly better than peak estimation. The average AUC values for all successive seasons stay close or even exceed the accuracy we have seen in the 2009 pandemic in onset estimation. The individual AUC values present a similar picture, although the trends inside epidemic seasons differ somewhat from those we have seen on Figure 3, with accuracy slowly decreasing as the outbreak progresses but stabilizing well above 0.8. In contrast, peak estimation performs much worse during most seasons compared with the 2009 pandemic. As in onset estimation, the more detailed results show a decreasing trend, but here stabilizing near 0.7, showing poor accuracy for peak estimation in the successive seasons.

**Fig. 7.**
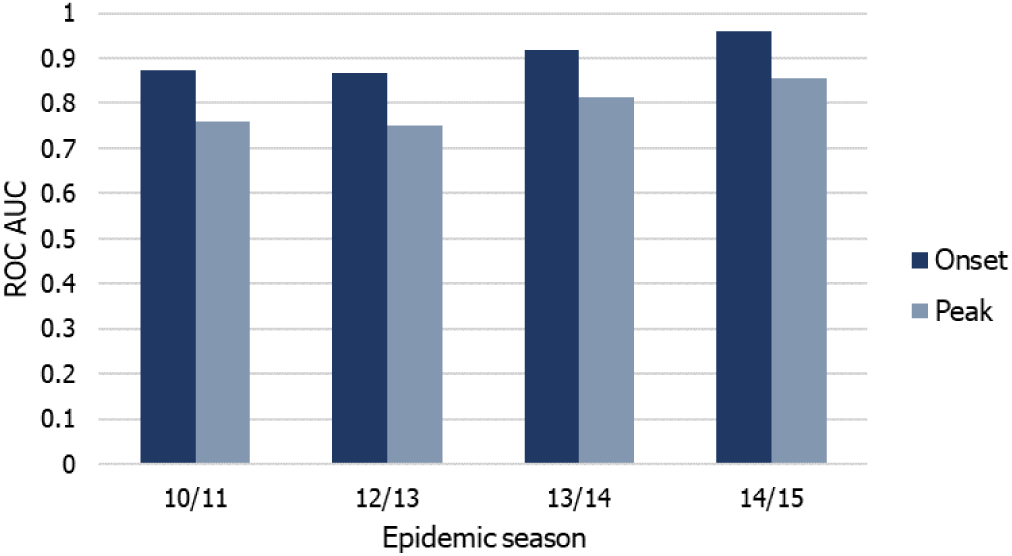
Predictive accuracy in onset and peak estimation for epidemic seasons 10/11, 12/13, 13/14, 14/15.

**Fig. 8.**
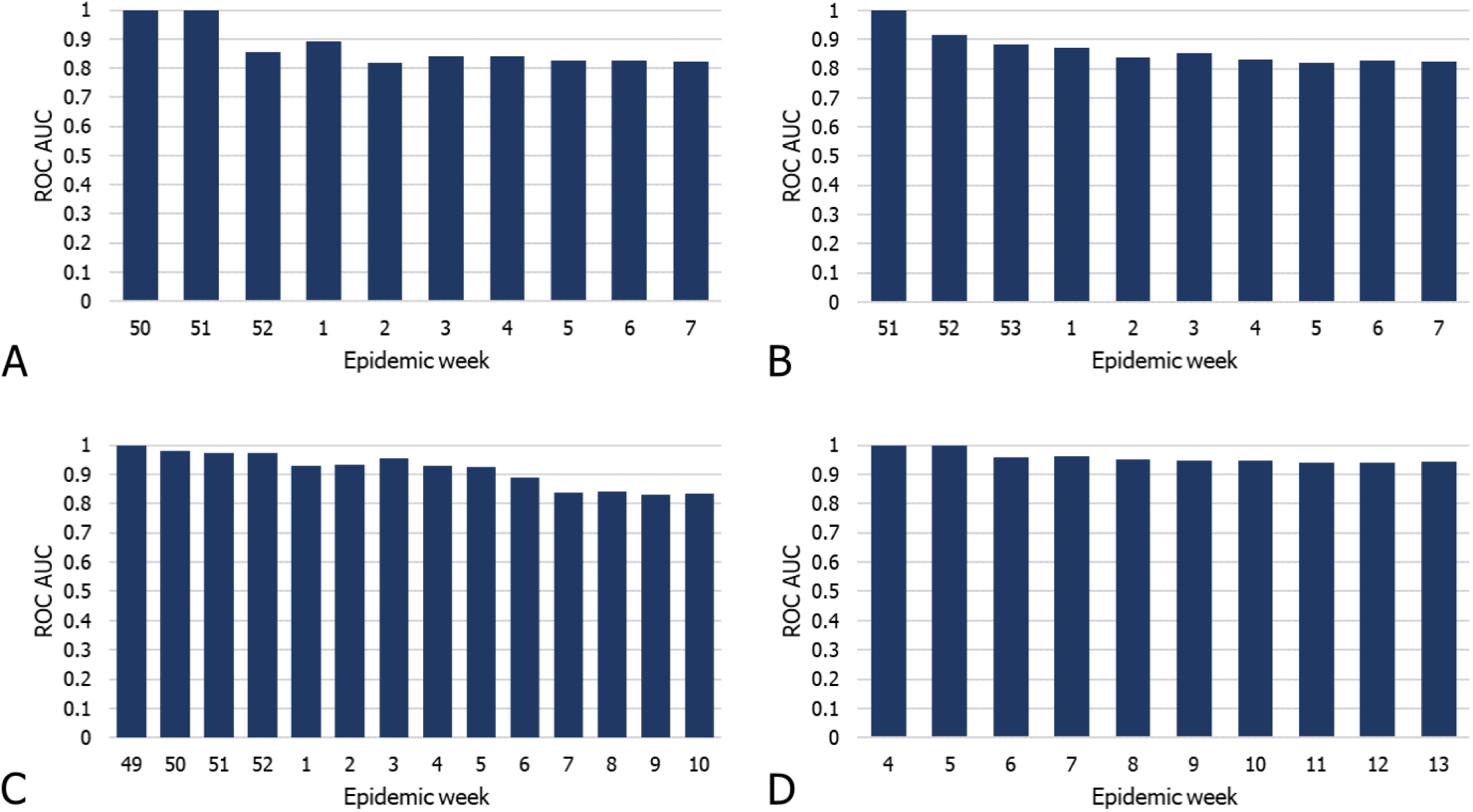
Predictive accuracy in onset estimation. A. Epidemic season 10/11. B. Epidemic season 12/13. C. Epidemic season 13/14. D. Epidemic season 14/15.

The predictive accuracy is greater in the 13/14 and 14/15 seasons than in the first two. While the weakening immunity granted to the population by the 09/10 pandemic may explain this trend, our results do not provide conclusive evidence to confirm this hypothesis.

We conclude that the onset prediction accuracy of the model is good, close to or even exceeding the accuracy of the 2009 pandemic itself. Predicting the timing of the epidemic peak is a much more difficult task for our model. Our results indicate that time of onset is more associated with structural factors, while peak time is more associated with environmental factors, making it more variable over seasons and thus less predictable. The predictive ability of both types of estimations improve in later seasons, and one possible explanation for this trend is the presence of immunity to the disease. Taking this factor into account in future work may improve the performance of our model.

## V. Conclusion

According to our findings, the spreading of H1N1 in Sweden was mainly driven by large population centers and the presence and size of outbreaks in neighboring municipalities. We find notable differences in the other risk factors contributing to the epidemic onset and peak. Confirming existing observations [6], [9] travel affects the spreading process, but it is less important than other factors. However, this might be the the result of the relatively small geographical scope of the study and the well-developed infrastructure of the country. According to our results, mean temperature plays a critical role in the timing of the epidemic peak, while it contributes only slightly to the onset of the outbreak. This result supports the narrative that meteorological factors aggravate existing outbreaks, driving them to a higher intensity. The only other socio-economic indicator that contributes noticeably to our model is the number of children per household, confirming the observations in [27]. However, like temperature, this indicator influences the epidemic peak much more than the onset of the outbreak. The effect of the rest of the socio-economic factors, income and social aid, is close to zero, likely because Sweden has one of the lowest income inequalities in the world.

We also showed, that while our model is constructed based on the 2009 pandemic, we can make accurate predictions on the timing of the selected epidemic events in the following seasons. Therefore, the model proposed in this paper can be used as a real-time decision support tool advising on resource allocation and surveillance. Furthermore, while our study only considers H1N1 spreading, it can be adapted to model other influenza strains or respiratory infections with a similar transmission mechanism.

## Data Availability

Data available on request from the authors

## Acknowledgment

Andras Bota was supported by the Olle Engkvist Byggmästare Foundation. Martin Rosvall was supported by the Swedish Research Council, grant 2016-00796.

